# Air and surface sampling for monkeypox virus in UK hospitals

**DOI:** 10.1101/2022.07.21.22277864

**Authors:** Susan Gould, Barry Atkinson, Okechukwu Onianwa, Antony Spencer, Jenna Furneaux, James Grieves, Caroline Taylor, Iain Milligan, Allan Bennett, Tom Fletcher, Jake Dunning, NHS England Airborne HCID Network

## Abstract

**Background:** An unprecedented outbreak of monkeypox virus (MPXV) infections in non-endemic countries has been recognised since 12 May 2022. More than 6000 cases have been identified globally with more than 1500 in the UK by July 2022. Transmission of MPXV is believed to be predominantly through direct contact with lesions or infected body fluids, with possible involvement of fomites and large respiratory droplets. Importantly, a case of monkeypox in a UK healthcare worker in 2018 was suspected to be due to virus exposure while changing bedding.

**Methods:** We investigated environmental contamination with MPXV from infected patients admitted to isolation rooms in the UK, to inform infection prevention and control measures. Surface swabs of high-touch areas in isolation rooms, of healthcare worker personal protective equipment (PPE) in doffing areas, and from air samples collected before and during bedding change were analysed using MPXV qPCR to assess contamination levels. Virus isolation was performed to confirm presence of infectious virus in key positive samples.

**Findings:** We identified widespread surface contamination (66 positive out of 73 samples) in occupied patient rooms (MPXV DNA Ct values 24·7-38·6), on healthcare worker personal protective equipment after use, and in doffing areas (Ct 26·3-34·3). Five out of fifteen air samples taken were positive. Significantly, three of four air samples collected during a bed linen change in one patient’s room were positive (Ct 32·7-35·8). Replication-competent virus was identified in two of four samples selected for viral isolation, including from air samples collected during the bed linen change.

**Interpretation:** These data demonstrate significant contamination in isolation facilities and potential for aerosolisation of MPXV during specific activities. PPE contamination was observed after clinical contact and changing of bed linen. Additionally, contamination of hard surfaces in doffing areas supports the importance of cleaning protocols, PPE use and doffing procedures.

**Funding:** No funding source for this study

## Introduction

An unprecedented number of cases of monkeypox have been confirmed outside endemic areas of West and Central Africa since 12 May 2022. As of the 5^th^ July, >6900 infections have been reported by more than 40 non-endemic countries; up to 7^th^ July, 1552 cases were reported by the UK.^1,2^

Monkeypox virus (MPXV) is an enveloped double stranded DNA virus classified within the *Orthopoxvirus* genus of the *Poxviridae* family. MPXV infection causes a clinical illness that is typically milder than smallpox, consisting of an influenza-like prodrome followed by a distinctive vesiculo-pustular rash. Lymphadenopathy typically occurs in monkeypox but not in smallpox.^3^ Mortality in monkeypox is thought to be between one and ten percent, influenced by clade and patient characteristics; as of 22 June 2022, only one death had been reported from non-endemic countries experiencing outbreaks during the current outbreak.^4^

Cases increased in endemic countries following the cessation of the smallpox vaccination program, and there have been concerns about potential increase in cases in travellers over the past decade.^5,6^ Primary cases arise from contact with animal reservoirs, and rodents are thought to have an important role, although further research is needed.^7^ Sustained human-to-human transmission has not been reported, and the secondary household attack rate in endemic settings has been reported to be between 0-10.2% in the majority of cases, and as high as 50% in one outbreak.^8^ Transmission to secondary cases is believed to be predominately via direct contact with body fluids or lesions, respiratory droplets and fomites. Infection by inhalation of high tire aerosolised Central African clade MPXV has been demonstrated in non-human primates, raising the possibility of potential aerosol transmission between humans, although existing epidemiological investigations suggest long-range aerosol transmission does not occur.^9,10^

Orthopox viruses are stable in the environment and can remain viable in aerosols for up to 90 hours.^11,12^ A hospital worker in the UK who developed monkeypox was thought to have been exposed to virus while changing the bedding used by a patient with monkeypox, before the diagnosis had been considered and appropriate infection control measures instigated.^13^ Widespread surface contamination in hospital rooms occupied by two patients with MPXV infection has recently been reported.^14^

The outbreaks occurring in higher resource settings in 2022 provide opportunities to investigate the extent of environmental contamination in symptomatic patient rooms within the airborne isolation units of NHS England’s High Consequence Infectious Diseases (HCID) Network, to inform practice around isolation, personal protective equipment, decontamination protocols and public health management of community exposures. There is also an opportunity to investigate whether aerosol transmission risks occur, and whether certain activities – such as changing bedding – increase the risk of exposure.

## Methods

### Sampling of patient rooms

Hospitalised adult patients with confirmed monkeypox and active skin lesions were identified, and verbal consent obtained to sample the air and environment within isolation rooms at the Royal Free Hospital. Air and surface sampling was performed in four positive pressure ventilated lobby (PPVL)^15^ single-occupancy respiratory isolation rooms (Room A, C, D and E including the negative pressure ensuite bathroom and positive pressure ventilated anterooms) in addition to the anterooms for three PPVL respiratory isolation rooms (Rooms A, B and C). The number of air changes per hour (ACH) greater than or equal to 10 in patient rooms and the average pressure differential between negative pressure areas (bedroom and bathroom) and other areas is maintained at 8 Pa or higher. Sampling of Room A and the anteroom of room C was performed twice with different occupants; these rooms were decontaminated using vapourised hydrogen peroxide and manual cleaning of surfaces with sodium hypochlorite solution between occupancies. Clinicians provided clinical data relevant to interpreting the results of environmental sampling e.g. recent virology results, date of onset of symptoms and date of admission.

Surface sampling using Copan UTM® swabs targeted high-touch areas, an air vent above the door leading to the ensuite bathroom, and a potential deposition area unlikely to have been directly touched by patient. Air sampling using the MD8 Airport (Sartorius, with gelatine filters; flow rate = 50L/min for ten mins) was performed before and during change of bed linen for the first visit to room A, all subsequent air sampling in rooms was for five minutes (flow rate=50 L/min). Air samplers were sited near to the bed (height ∼1 m, distance from patient bed ∼1 m) and further away (height ∼2 m, distance from bed >1.5 m). Wearable samplers (SKC, with gelatine filters; flow rate 4 L/min for ten mins) were utilised on the first visit to room A including by the healthcare worker (HCW) performing the linen change.

Minimal anonymised epidemiological and clinical data was provided by treating clinical team including date of admission, date of onset of illness, most recent virology results and whether the patient has received tecovirimat. All patients provided written informed consent for the ISARIC Clinical Characterisation Protocol (ref) that includes air and environmental sampling. The study was undertaken as an Urgent Public Health Investigation with UK HSA Research Ethics and Governance of Public Health Practice Group (REGG) approval.

### Sampling of PPE

Staff entering an isolation room wear disposable single-use PPE (surgical gown, plastic apron, double nitrile gloves, FFP3 respirator, hair cover, and autoclavable plastic clogs) which is donned prior to entering the anteroom. To exit a patient room, staff enter the anteroom which is demarcated into toxic (close to patient room) and non-toxic (close to exit to the corridor) using tape on the floor. PPE doffing protocols within the anteroom are designed to limit contamination of the non-toxic area with potentially contaminated items entering waste-streams located in the toxic area. Staff transfer into clean clogs after all PPE apart from scrubs has been remove as they transition to the non-toxic part of the anteroom before a final hand wash prior to exit. This process is monitored by a buddy to ensure all parts are executed in the correct order.

Prior to the removal of PPE in the anteroom, swabs were taken of the front of the gown, gloves and visor of HCWs who had either had clinical contact (rooms B and C) or changed the bedding (room A). Swabs were taken of the floor in the doffing area immediately after PPE removal in each case. Air samples were taken simultaneously in the anteroom and in the adjacent corridor prior to, and during, the doffing procedure using the MD8 Airport with gelatine filters (50 L/min for five minutes) in rooms A-C.

### Sample processing

For surface samples, 140 µL of UTM was inactivated using Buffer AVL (Qiagen) with nucleic acid using the Viral RNA Mini Kit (Qiagen) in accordance with the manufacturer’s instructions. For air samples, gelatine filter were dissolved in 20 mL of warmed MEM media (Gibco) for MD8 filters, or 5ml for personal sampler gelatine filters, with 140 µL then used for inactivation and extraction as described above. Analysis of extracted nucleic acid for the presence of MPXV DNA was performed using a published assay with minor modifications to conform with local standardised diagnostic processes.^16^

### Viral Isolation

Four key samples were selected for viral isolation. 0.5 mL of UTM (swab samples) or MEM media containing dissolved gelatine filter (air sample) was added to a 70% confluent monolayer of Vero C1008 cells (ECACC 85020206) in a T-25cm2 non-vented tissue culture flasks (Corning) and incubated for 1hr at 37°C. After 1 hr, the inoculum was removed and washed with sterile PBS before addition of 5mLviral culture medium consisting of 1 x MEM + GlutaMAX™ supplemented with 5% heat-inactivated FBS, 25mM HEPES, and 4x antibiotic-antimycotic solution (all Gibco). A negative control flask was also prepared by the same method using 0.5 mL of MEM as the inoculum. All flasks were incubated at 37°C and monitored for cytopathic effect (CPE) using a phase contrast inverted light microscope. 140 µL timepoints were collected every 48-72 hours to monitor for a change in detectable DNA via qPCR. Over-confluent cell monolayers (five days post infection) that did not display viral CPE were passaged by inoculating supernatant onto fresh sub-confluent cells providing continuous assessment for CPE for 10 days.

## Results

MPXV DNA was detected in 56 of 60 (93%) surface swab samples obtained within the patients’ bedrooms and bathrooms, with Ct values between 24·7 and 37·4 (Table 1). All patients had monkeypox lesions present on multiple areas of their bodies at the time of environmental sampling. The detections included samples from areas unlikely to have been directly touched by patient, such as the air vent above the door between the bedroom and the bathroom, suggesting non-contact contamination possibly via respiratory droplets or re-aerosolisation from activities such as changing bed linen.

**Table 1:** Clinical characteristics and results of surface and air environmental sampling in patients’ rooms. Details of environmental sampling performed in five patient rooms at the Royal Free Hospital May-June 2022. P1/P2 = Rooms A was sampled on two occasions with different patients occupying this room on each visit; rooms were decontaminated every 12hrs during occupancy using 5,000ppm available chlorine sodium hypochlorite on all surfaces and 10,000 ppm available chlorine sodium hypochlorite for toilet, shower, wash basins and floors with a full room clean after patient discharge, followed by decontamination using vapourised hydrogen peroxide. *Denotes occupant of this room was the first patient admitted into this room with monkeypox Ct = qPCR crossing threshold value of MPXV DNA detected. NA = Not applicable (sample not taken for this room). NR = Tecovirimat not received.

|  | **Room A**<br>**(P1)\*** | **Room A**<br>**(P2)** | **Room C**<br>**(P2)** | **Room D\*** | **Room E\*** |
| --- | --- | --- | --- | --- | --- |
| **Background information** |  |  |  |  |  |
| *Date of sampling* | *24<sup>th</sup> May* | *17<sup>th</sup> June* | *16<sup>th</sup> June* | *17<sup>th</sup> June* | *16<sup>th</sup> June* |
| *Days since onset* | 9 | 30 | 6 | 26 | 7 |
| *Days since admission* | 2 | 7 | 7 | 18 | 3 |
| *Throat Ct at admission* | 27 | Negative | 22 | 37 | 30 |
| *Lesion Ct at admission* | 22 | 23 | 28 | 23 | 31 |
| *Plasma Ct at admission* | 32 | 34 | 35 | Negative | 31 |
| *Days on tecovirimat* | 2 | 4 | NR | NR | 3 |
| **Patient Room Ct values** |  |  |  |  |  |
| Floor | NA | 26.9 | 30.9 | 34.9 | 32.5 |
| Call button | 27.5 | 29.4 | 32.4 | Negative | 26.1 |
| Light switch | 24.7 | 31.6 | 34.5 | 36.3 | 30.2 |
| TV remote control | 25.0 | 28.9 | 37.4 | 32.2 | 28.2 |
| Observation machine | 26.4 | NA | NA | NA | NA |
| Tap handle (bedroom) | 32.4 | 34.2 | 35.6 | 36.7 | 27.1 |
| Window ledge | 28.8 | 29.7 | Negative | 35.6 | 35.5 |
| Chair – arm rest | 29.9 | 33.5 | 33.8 | 31.6 | 24.9 |
| Door handle (room to bathroom) | 26.7 | 33.3 | 32.6 | Negative | 28.1 |
| **Bathroom Ct values** |  |  |  |  |  |
| Vent/grille (room to bathroom) | 26.4 | 25.9 | 27.9 | 33.3 | 33.6 |
| Toilet flush handle | 28.7 | 32.6 | 31.8 | 34.8 | 26.4 |
| Shower handle | 28.8 | 33.5 | 34.0 | 33.8 | 32.7 |
| Tap handle (bathroom) | 29.2 | 29.3 | 32.8 | Negative | 25.9 |
| **Anteroom Ct values** |  |  |  |  |  |
| Floor: toxic side | 26.3 | 28.7 | 33.2 | 32.9 | 30.6 |
| Floor: non-toxic side | NA | 33.6 | Negative | 36.8 | 36.8 |
| **Ward Ct values** |  |  |  |  |  |
| Corridor | NA | Negative | 37.5 | Negative | 36.7 |
| **Air sampling Ct values** |  |  |  |  |  |
| Pre bed change near | Negative | Negative | Negative | Negative | Negative |
| Pre bed change far | 36.2 | 36.5 | Negative | Negative | Negative |
| During bed change near | 32.7 | 36.2 | Negative | Negative | Negative |
| During bed change far | 35.8 | Negative | Negative | Negative | Negative |

Following doffing of PPE by hospital staff, MPXV DNA was detected on the floor of the anteroom where doffing took place (Table 2). MPXV DNA was detected (Ct 38·2) in one air sample taken in the anteroom prior to doffing, but not in other air samples taken before and during doffing in anterooms used for doffing, nor in the corridor outside the anterooms. The volume of air filtered in air sample collected in the anteroom and corridor was 250 litres (50L/min for five minutes). MPXV DNA was not detected in samples from any wearable air sampler. By contrast, and of importance, five of eight air samples collected over ten minutes by the large volume air sampler (flow rate 50L/min) in Room A contained MPXV DNA; these samples were collected in two sets approximately three weeks apart with different patients in this room at the time each sampling was performed. These results include samples taken further away from the patient (>1·5m), and observation of a lower Ct value for the sample obtained during the bedding change compared to the Ct values for samples collected before the bedding change (Figure 1). MPXV DNA was not detected in air samples collected over five minutes at the same distances prior to and during bedding changes in rooms C-E.

**Table 2:**
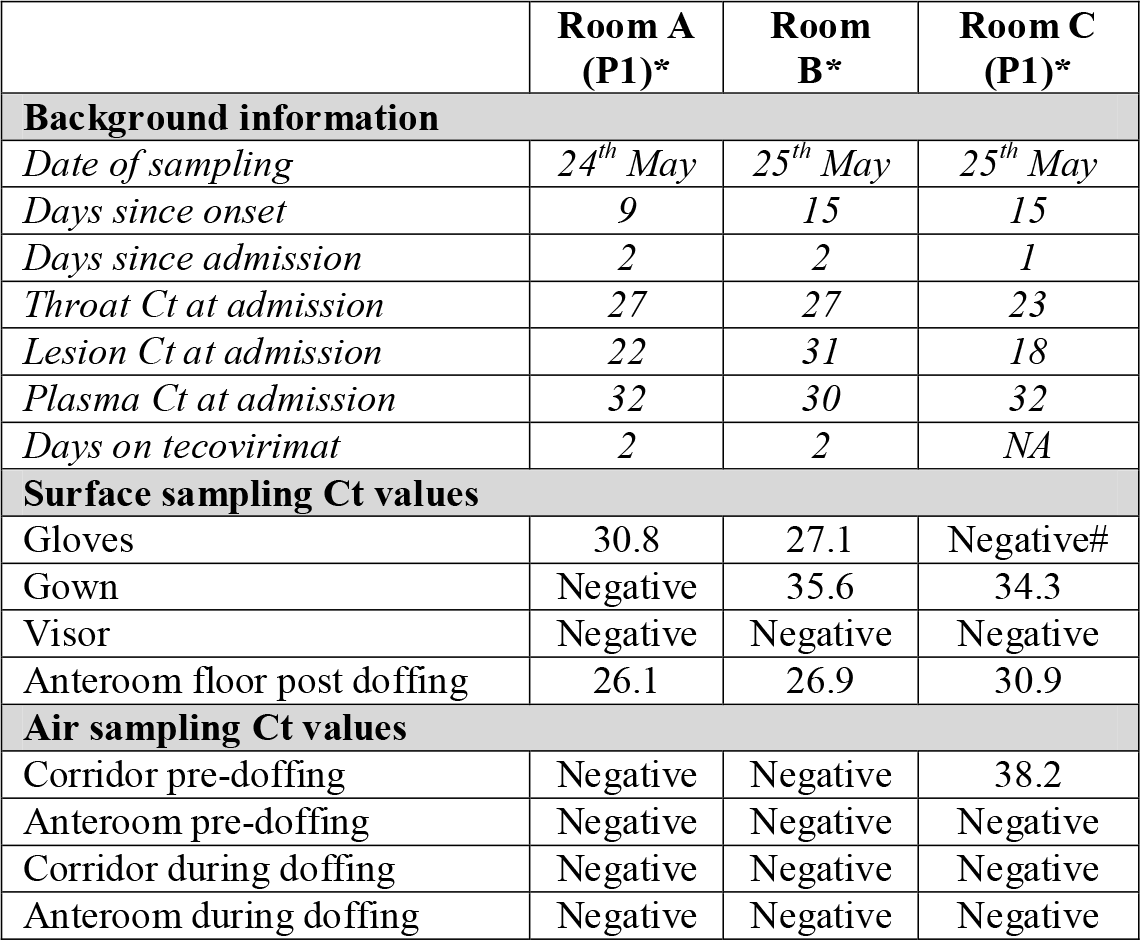
Clinical characteristics and results of sampling around doffing procedure. Details of environmental sampling performed around doffing procedure at the Royal Free Hospital May 2022. P1/P2 = Rooms A and C were both sampled on two occasions with different patients occupying these rooms on each visit; rooms were decontaminated every 12hrs using 5,000ppm available chlorine sodium hypochlorite during occupancy with a full room clean with 5,000ppm available chlorine sodium hypochlorite after patient discharge, followed by decontamination using vapourised hydrogen peroxide. *Denotes occupant of this room was the first patient admitted into this room with monkeypox. #This negative result is likely due to sampling of the palmar surface only rather than palmar surface and fingertips. Ct = qPCR crossing threshold value of MPXV DNA detected.

|  | Room A (P1)* | Room B* | Room C (P1)* |
| --- | --- | --- | --- |
| **Background information** |  |  |  |
| *Date of sampling* | *24<sup>th</sup> May* | *25<sup>th</sup> May* | *25<sup>th</sup> May* |
| *Days since onset* | 9 | 15 | 15 |
| *Days since admission* | 2 | 2 | 1 |
| *Throat Ct at admission* | 27 | 27 | 23 |
| *Lesion Ct at admission* | 22 | 31 | 18 |
| *Plasma Ct at admission* | 32 | 30 | 32 |
| *Days on tecovirimat* | 2 | 2 | NA |
| **Surface sampling Ct values** |  |  |  |
| Gloves | 30.8 | 27.1 | Negative# |
| Gown | Negative | 35.6 | 34.3 |
| Visor | Negative | Negative | Negative |
| Anteroom floor post doffing | 26.1 | 26.9 | 30.9 |
| **Air sampling Ct values** |  |  |  |
| Corridor pre-doffing | Negative | Negative | 38.2 |
| Anteroom pre-doffing | Negative | Negative | Negative |
| Corridor during doffing | Negative | Negative | Negative |
| Anteroom during doffing | Negative | Negative | Negative |

**Figure 1:**
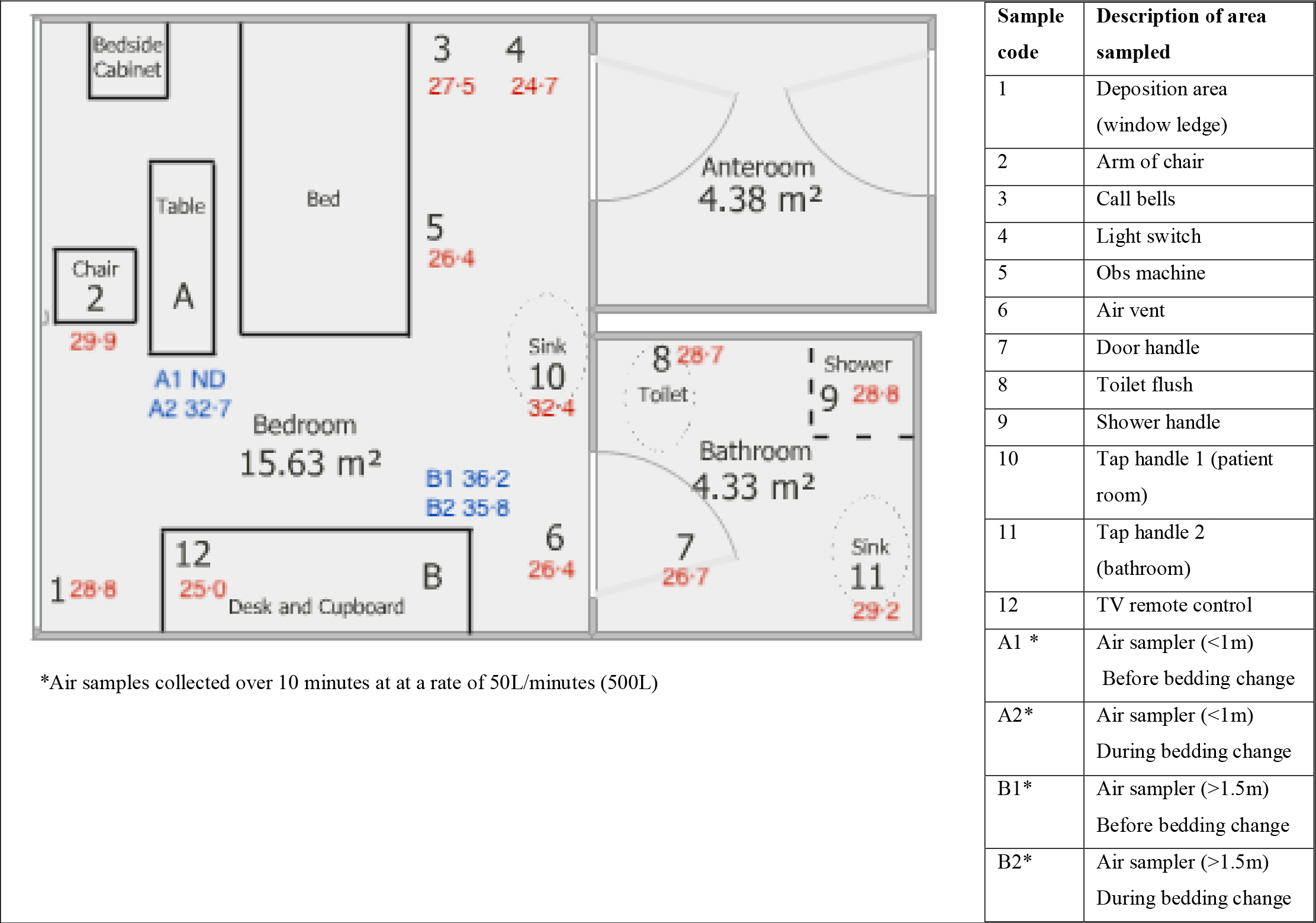
Plan of Room A representing sites of sampling and MPXV PCR Ct values.

Previous work suggests that MPXV is likely to be culturable from samples with a Ct value below 30 for this particular reference laboratory PCR assay.^17^ Virus isolation was attempted for four samples: Room A light switch (original Ct 24.7), Room A anteroom floor after PPE doffing (original Ct 26.3), Room B anteroom floor after PPE doffing (original Ct 26.9) and near bed air sample during bed linen change in Room A (original Ct 32.7). While no flask showed marked CPE during the experiment, viral DNA replication was witnessed for both Room B anteroom floor swab sample and Room A bed change air sample with Ct values recorded as >31.0 cycles for all timepoints between Day 0 and Day 5, but with Ct values <16.0 for Day 10, indicating at least 100,000 times more detectable DNA between the Day 5 and Day 7 timepoint (Table 3). Only very subtle CPE was witnessed in the Room B sample with no obvious CPE is the Room A air sample. The other two swab samples, and the negative control, showed no increase in detectable DNA during the ten-day isolation attempt.

**Table 3:** MPXV Ct values at specified timepoints from viral isolation cultures. qPCR crossing threshold values from viral isolation cultures of environmental samples. Ct = Crossing threshold value; NA = Not applicable; ND = Not detected; P0/P1 = passage 0/1.

|  | No infection control | Room B Anteroom floor | Room A (P1) Bed change | Room A (P1) Anteroom floor | Room A (P1) Light switch |
| --- | --- | --- | --- | --- | --- |
| **Environmental sample Ct** | NA | 26.9 | 32.7 | 26.3 | 24.7 |
| **Day 0 P0** | ND | 32.2 | ND | 33.5 | 32.2 |
| **Day 3 P0** | ND | ND | ND | 33.6 | 33.1 |
| **Day 5 P0** | ND | 31.4 | 35.5 | 36.3 | 38.1 |
| **Day 7 P0** | ND | 22.4 | 27.6 | ND | 35.6 |
| **Day 7 P1 (5+2)** | ND | 39.5 | ND | ND | ND |
| **Day 10 P1 (5+5)** | ND | 14.9 | 17.5 | 36.2 | 36.5 |
| **Cultured MPXV?** | No | Yes | Yes | No | No |

## Discussion

We identified widespread MPXV DNA contamination of the environment in respiratory isolation rooms occupied by infected symptomatic individuals. MPXV PCR Ct values obtained from these samples are within the range previously shown to be associated with recovery of infection-competent MPXV. We found evidence of replication-competent virus from two samples including from an air sample collected during a bed linen change. Detection of infectious MPXV in air samples collected during a bed linen change highlights the importance of suitable respiratory protection equipment when performing activities that may re-aerosolise infectious material within contaminated environments. The variability in the frequency of detection and the Ct values observed in surface samples from different patient rooms may be due to individual patient factors, the point during patient infection that environmental sampling was performed, staff or patient behaviour, and the frequency of cleaning.

Significant contamination of PPE was also found following clinical contact with a patient (rooms B, C) or bed linen (room A). MPXV DNA was detected on the glove samples where swabbing included both palmar surface and fingertips, and not detected on a sample collected only from the palmar surface. The detection of MPXV DNA with relatively low Ct values in hard-surface samples from the doffing environment reinforces the importance of surface cleaning protocols, the use of appropriate PPE, and robust doffing procedures to maintain the safety of staff and avoid potential onward transmission. Although our findings are specific to sampling in a specialist healthcare environment and sampling within occupied rooms was limited to a small number of patients, the environmental contamination findings may be relevant to public health measures for other spaces and settings where individuals with monkeypox spend prolonged periods, such as residential bedrooms and bathrooms. Further investigation is required into the contamination of areas occupied for shorter periods of time, such as outpatient clinics, and also healthcare spaces that do not have mechanical negative pressure ventilation. Previous investigations of surface contamination in a domestic setting and in two hospital rooms occupied by infected symptomatic individuals have demonstrated a high frequency of MPX viral DNA detection and isolation of virus.^14,17^

To the best of our knowledge, this is the first time that detection of MPXV (DNA and virus by isolation) in environmental air samples from healthcare settings has been reported, for any clade of MPXV. Detection of MPXV DNA in air samples collected at distances of greater than 1·5m from the patient and at a height of nearly 2m supports the theory that MPXV can be present in either aerosols, suspended skin particles or dust containing virus, and not only in large respiratory droplets that fall to the ground within 1 to 1·5m of an infected individual. Low flow-rate wearable button samplers provided negative samples but were only deployed for under ten minutes, which may be an insufficient sampling time (<40L of air sampled). Our findings support recommendations for healthcare workers interacting with patients with confirmed MPXV infection to use suitable PPE, including respiratory protective equipment, as well as other IPC measures designed to limit exposure to pathogens that may become aerosolised in hospital inpatient settings.

## Data Availability

All data produced in the present study are available upon reasonable request to the authors

## Acknowledgments

The authors gratefully acknowledge the medical, nursing and multidisciplinary staff supporting the Airborne HCID Treatment Centres at the Royal Free London NHS Foundation Trust and the Royal Liverpool University Hospital, the NHS England Airborne HCID Network, the UK Health Security Agency, and the Rare and Imported Pathogens Laboratory at UKHSA Porton Down. The authors also acknowledge the assistance of Ian Nicholls and Wilhemina D’Costa (both UKHSA) for assisting with nucleic acid extractions, and from Sian Summers (UKHSA) for providing cell culture flasks.

## Notes

**Sources of funding:** SG, JD and TF are supported by the National Institute for Health Research (NIHR) Health Protection Research Unit in Emerging and Zoonotic Infections. SG is supported by the Research, Evidence and Development Initiative (READ-It). READ-It (project number 300342-104) is funded by UK aid from the UK government; however, the views expressed do not necessarily reflect the UK government’s official policies

### Competing Interest Statement

The authors have declared no competing interest.

### Funding Statement

This study did not receive any funding

### Author Declarations

The investigations performed were a component of the urgent public health investigation performed as part of UKHSA’s public health incident response to cases of a high consequence infectious disease in the UK. UKHSA is the national health security agency for England and an executive agency of the UK Government’s Department of Health and Social Care. The study protocol was subject to internal review by the Research Ethics and Governance Group, which is the UKHSA Research Ethics Committee, and was granted full approval.

