## Appendix 1 for "Air and surface sampling for monkeypox virus in UK hospitals"

Appendix 1: NHS England Airborne HCID Network

Jake Dunning,^1^ Nicholas Price,^2^ Michael Beadsworth,^3^ Matthias Schmid,^4^ Marieke Emonts,^4^

Anne Tunbridge,^5^ David Porter,^6^ Jonathan Cohen,^7^ Elizabeth Whittaker^8^ and Ruchi Sinha,^8^

1. Royal Free London NHS Foundation Trust, London
2. Guys and St Thomas’ NHS Foundation Trust, London
3. Royal Liverpool University Hospital, Royal Liverpool University Hospitals Foundation Trust, Liverpool
4. Newcastle Royal Victoria Infirmary, Newcastle Hospitals NHS Foundation Trust, Newcastle
5. Royal Hallamshire Hospital, Sheffield Teaching Hospitals NHS Foundation Trust, Sheffield
6. Alder Hey Children’s Hospital, Liverpool
7. Evelina London Children’s Hospital, London
8. Imperial College Healthcare at St Mary’s Hospital, London
